# Brain injury biomarkers in major and simple neurocognitive psychosis: association with tryptophan catabolites

**DOI:** 10.1101/2025.01.22.25320986

**Authors:** Hussein Kadhem Al-Hakeim, Ameer Abdul Razzaq Al-Issa, Mengqi Niu, Yingqian Zhang, Michael Maes

**Affiliations:** Sichuan Provincial Center for Mental Health, Sichuan Provincial People’s Hospital, School of Medicine, University of Electronic Science and Technology of China, Chengdu 610072, China; Key Laboratory of Psychosomatic Medicine, Chinese Academy of Medical Sciences, Chengdu, 610072, China; Department of Chemistry, Faculty of Science, University of Kufa, Najaf, Iraq; Department of Psychiatry, Faculty of Medicine, Chulalongkorn University, Bangkok, Thailand; Cognitive Impairment and Dementia Research Unit, Faculty of Medicine, Chulalongkorn University, Bangkok, Thailand; Cognitive Fitness and Biopsychological Technology Research Unit, Faculty of Medicine Chulalongkorn University, Bangkok, 10330, Thailand, Bangkok, 10330, Thailand; Department of Psychiatry, Medical University of Plovdiv, Plovdiv, Bulgaria; Research Institute, Medical University of Plovdiv, Plovdiv, Bulgaria; Kyung Hee University, 26 Kyungheedae-ro, Dongdaemun-gu, Seoul 02447, Korea; Research and Innovation Program for the Development of MU - PLOVDIV–(SRIPD-MUP)”, Creation of a network of research higher schools, National plan for recovery and sustainability, European Union – NextGenerationEU

**Keywords:** neurotoxicity, schizophrenia, neuroimmune, oxidative stress, inflammation, biomarkers

## Abstract

**Background:** Schizophrenia is categorized into qualitatively distinct classes, i.e. major (MNP) and simple (SNP) neurocognitive psychosis. MNP is accompanied by more severe neurocognitive deficits and symptomatology, activated immune-inflammatory and oxidative stress pathways, and induction of the tryptophan catabolite (TRYCAT) pathway with increased quinolinic acid (QA) and lowered kynurenic acid (KA) levels.

**Aims:** To examine whether MNP and increased QA levels are associated with increased brain injury markers, including S100 calcium-binding protein B (S100B), neuroepithelial stem cell protein (Nestin), neuron-specific enolase (NSE), phosphorylated tau217 (pTau217), and glial fibrillary acidic protein (GFAP).

**Methods:** This case-control study included 52 MNP subjects, 68 SNP subjects, and 60 healthy controls and assessed the above brain injury biomarkers and TRYCATs.

**Results:** NSE and GFAP were significantly higher in MNP than in SNP, and in both MNP or SNP than in controls. Serum S100B levels were substantially higher in MNP than in controls and SNP. The results indicate injuries to neurofilaments in MNP and SBP, and that MNP is additionally characterized by damage to cell bodies, axons, glial cell projections, reduced neurogenesis and synaptic plasticity as compared with SNP. Increased QA levels and lowered KA predict increased pTau217, NSE and GFAP. The QA/KA ratio is the best predictor of these three brain injury markers

**Conclusions:** These findings validate the differentiation between the two distinct subclasses with MNP being characterized by more profound injuries to brain cells and structures as compared with SNP. Increases in peripheral QA levels may contribute to these brain injuries in MNP.

## Introduction

Despite a consensus on the clinical heterogeneity of chronic schizophrenia, no subclassification strategy has garnered general acceptance. Most methods for defining subgroups have depended solely on the course or kind of symptoms (Kremen et al., 2004, Joyce and Roiser, 2007, Jiang et al., 2024). However, in 2018, a new and cross-validated categorization was introduced utilizing unsupervised machine learning approaches (Kanchanatawan et al., 2018). The authors identified that schizophrenia can be categorized into two separate diagnostic groups: major neurocognitive psychosis (MNP) and simple neurocognitive psychosis (SNP) (Kanchanatawan et al., 2018b, Kanchanatawan et al., 2018a)(Maes, 2023b). MNP and SNP are two distinct categories based on the outcomes of neuropsychological tests, symptom profiles, and serum biomarker levels (Popov et al., 2024, Kanchanatawan et al., 2018a).

MNP is characterized by elevated negative symptoms, elevated PHEM (psychotic, hostility, excitation, and mannerism), and overall severity of schizophrenia (OSOS) scores in comparison to SNP (Maes, 2023a, Maes et al., 2020a, Sirivichayakul et al., 2019b, Almulla et al., 2021). Moreover, MNP as compared with SNP is accompanied by an activated immune-inflammatory response system (IRS), including M1 macrophage, T helper (Th)1, and Th17 immune profiles, increased oxidative stress, and increased translocation of Gram-negative bacteria or their lipopolysaccharides (LPS) (Maes et al., 2019b, Maes et al., 2021, Popov et al., 2024). Furthermore, MNP is characterized by lowered activities of the compensatory immunoregulatory system (CIRS), including lowered protective IgM-mediated autoimmune responses, and lowered antioxidant defenses (Maes, 2023a). It is crucial to note that the increased IRS activities and relative deficiencies in the CIRS pathways can account for a substantial portion of the variance in the scores of overall severity of schizophrenia (OSOS) (Maes et al., 2019a, Maes et al., 2019b). Lowered CIRS activities diminish the protection against the neurotoxic and apoptotic effects of some cytokines, oxidative stress, hypernitrosylation, and associated enzymes that may inflict brain cell injuries often leading to cell death (Roomruangwong et al., 2020; Manev et al., 1989). In light of this, it is essential to consider the newly machine learning-derived classification when doing research on biomarkers of schizophrenia.

There is currently evidence to suggest that schizophrenia is associated with aberrations in the tryptophan catabolite (TRYCAT) or kynurenine (KYN) pathway when contrasted with controls. For example, a recent study revealed that kynurenic acid (KA) and the KA/KYN ratio were markedly elevated in schizophrenia patients compared to healthy controls, after adjusting for age and sex (Zhou et al., 2022). Nevertheless, it appears that such differences in schizophrenia may be attributable to the increased TRYCAT levels in MNP versus SNP (Kanchanatawan et al., 2018a; Al-Hakeim et al., 2024). The former study observed that IgA responses were elevated in response to neurotoxic TRYCATs, including KYN, xanthurenic acid, and picolinic acid, while responses to TRYCATs that were generally more protective, such as anthranilic acid (AA) and KA, were reduced. In comparison to SNP patients and controls, the latter authors (Al-Hakeim et al., 2024) observed increased levels of serum AA and quinolinic acid (QA) in MNP versus SNP and controls, as well as lower levels of serum tryptophan, kynurenic acid (KA), and 3-OH-anthranilic acid (3HAA) in patients with MNP. The best biomarker of MNP was an increase in the QA/KA ratio. Thus, changes in serum TRYCAT levels further substantiate that MNP and SNP represent two distinct subgroups of schizophrenia from a biological perspective. A recent meta-analysis indicated that individuals with schizophrenia had significantly reduced peripheral blood TRP levels compared to controls (Almulla et al., 2022). Other studies found lower serum KYN and KA levels in schizophrenia (Yang et al., 2022) or lower serum KYN, XA, QA and PA levels in a combined patient group of schizophrenia and bipolar disorder (Skorobogatov et al., 2023).

These changes may have dire consequences since the TRYCAT pathway is mainly a protective, anti-oxidant and anti-inflammatory pathway (Al-hakeim et al., 2024). Consequently, decreased levels of some TRYCATs in MNP may adversely affect antioxidant and immunoregulatory functions, whereas increased QA production may have neurotoxic effects. Thus, elevated levels of QA may exacerbate breakdown of the blood-brain barrier (review: Al-Hakeim et al., 2024), which has been described in MNP (Maes et al., 2019b). The consequent infiltration of QA into the CNS may activate N-methyl-D-aspartate receptors (NMDARs) resulting in influx of calcium ions into the postsynaptic cell, excitotoxicity, and brain cell injuries (Neves et al., 2023; Zhang et al., 2019; González-Cota et al., 2024). Such effects may be potentiated by lowered levels of KA, which have been decsribed in MNP (Al-Hakeim et al., 2024). Furthermore, QA levels, especially when coupled with decreases in KA, may promote the production of reactive oxygen species during mitochondrial oxidative stress, leading to brain cell injuries (Yu et al., 2016; Cao et al., 2021; Kindler et al., 2020).

Damage to brain cells, including neurons and astroglia, is accompanied by increased release of neuron or astroglia damage markers (Wang et al., 2015, Silvestro et al., 2024, Huibregtse et al., 2021). Important damage markers include S100 calcium-binding protein B (S100B) (das Neves et al., 2021), neuroepithelial stem cell protein (Nestin) (Bornstein et al., 2020), neuron-specific enolase (NES) (Palumbo et al., 2008), phosphorylated tau217 (pTau217) (Palmqvist et al., 2020), and glial fibrillary acidic protein (GFAP) (Brenner, 2014). These biomarkers are measurable in serum and indicate damage to brain cells. Nonetheless, there are no studies that have examined these brain injury biomarkers to differentiate between MNP and SNP relative to healthy controls, and there are no studies whether alterations in the TRYCAT pathway (e.g. increased QA levels) are associated with increased brain injury biomarkers.

Hence, the present study examines these brain injury markers in MNP versus SNP versus controls and examines whether the increase in serum levels of biomarkers is associated with the effects of neurotoxic TRYCATs like QA and / or with lowered levels of more protective TRYCATs such as KA and AA. The specific hypotheses are that MNP patients show increased levels of the damage biomarkers and that the damage biomarkers are positively associated with QA levels and inversely with the protective TRYCATs.

## Subjects and Methods

### Participants

In addition to sixty healthy controls, the current study recruited 120 patients diagnosed with schizophrenia subtypes MNP or SNP. The Ibn-Rushd Training Hospital for Psychiatric Medicine in Baghdad, Iraq, recruited all participants from January 2024 to April 2024. The year preceding the investigation, all patients with schizophrenia were in a stable phase of their illness and had not yet experienced any acute episodes. Patients were diagnosed with “schizophrenia” in accordance with the DSM-IV-TR criteria. Finally, we included patients with schizophrenia who met the diagnostic criteria of MNP as outlined by Kanchanatawan et al. (2018a) (Kanchanatawan et al., 2018a). Patients who did not satisfy these criteria were classified as SNP. Al-Hakeim et al. (2024) have published all the details of the samples that were previously employed to investigate the TRYCATs in schizophrenia. The research conducted in this study has been approved by the ethics committee (IRB) of the College of Science, University of Kufa, Iraq (T4220/2023). This approval is in accordance with the International Guideline for Human Research Protection, as mandated by the Declaration of Helsinki.

### Measurements

#### Clinical assessments

In order to collect clinical and socio-demographic data from both patients and controls, a senior psychiatrist who specializes in schizophrenia, conducted a semi-structured interview. Using the DSM diagnostic criteria, the Mini-International Neuropsychiatric Interview (M.I.N.I.) was implemented to diagnose schizophrenia. The Arabic translation of the interview was validated in the Iraqi dialect. The same psychiatrist also assessed the Positive and Negative Syndrome Scale (PANSS) (Kay et al., 1987), the Scale for the Assessments of Negative Symptoms (SANS) (Andreasen, 1989), and the Brief Psychiatric Rating Scale (BPRS) (Overall and Gorham, 1962). As previously reported (Maes et al., 2020a, Sirivichayakul et al., 2019b, Sirivichayakul et al., 2019a), we employed z-unit weighted composite scores to calculate scores for the psychosis, hostility, excitation, and mannerism (PHEM) domains. The cumulative scores of the PANNS and SANS negative subscales served as measures for the severity of negative symptoms. We subsequently calculated the scores from the first initial principal component (PC) derived from psychosis, hostility, excitement, mannerism, the negative PANNS subdomain score, and the overall SANS score, as previously outlined (Al-Hakeim et al., 2024). This principal component analysis adheres to rigorous standards, including a KMO value exceeding 0.8 (specifically 0.933), a significant Bartlett’s test of sphericity (χ2=1534.446, df=15, p<0.001), an explained variance surpassing 50% (specifically 87.20%), and all loadings of the first principal component exceeding 0.7 (actually all are above 0.852).

#### Assays

All participants donated five millilitres of fasting blood in the morning (6.30–8.00 a.m.). After ten minutes, clotted blood samples were centrifuged at 1100 Xg for five minutes. Aliquoted serum into three Eppendorf tubes. In preparation for testing, the tubes were stored at -80 °C. We measured blood levels of human tryptophan (TRP), KYN, KA, AA, 3HAA, 3-hydroxy-kynurenine (3-HK), QA, S100B, pTau217, Nestin, NSE, and GFAP using commercially available ELISA kits from Nanjing Pars Biochem Co., Ltd. (Jiangsu, China). These ELISA kits are based on sandwich ELISA methods. All ELISA kits have intra-assay CVs below 10.0%. Samples with high analyte concentrations were diluted. The data on TRYCATs have been pre-published previously (Al-Hakeim et al., 2024). We calculated z unit-based composite scores reflecting a) the QA/KA ratio as explained previously, namely z QA – z KA (Al-Hakeim et al., 2024); and b) the combined effects of all 5 damage biomarkers as z S1008 + z pTau217 + z nestin + z NSE + z GFAP, labeled brain injury index (BII).

### Statistical analysis

A one-way analysis of variance was employed to compare scale variables among groups, whilst contingency tables (χ2 tests) were utilized to evaluate associations between ordinal variables. Pearson’s product-moment correlation, Spearman’s rank-order correlation coefficients, and partial correlation coefficients were employed to examine the associations among scale variables, subsequent to the adjustment for extraneous variables. Multiple regression analysis was employed to identify the biomarkers that predict the symptom domains by manual and automated stepwise methods (p-to-entry of 0.05 and p-to-remove of 0.06) and to evaluate the change in R^2^. Additionally, the analysis was assessed for collinearity (utilizing VIF and tolerance) and homoscedasticity (employing the White and Breusch-Pagan tests). Following the rejection of the latter, we employed heteroscedasticity-consistent standard error (SE) or robust SE estimations derived from the HC3 method. The associations among diagnostic groups (MNP, SNP, and controls) and BI biomarkers were examined utilizing multivariate and univariate GLM models that accounted for confounding variables (age, gender, education, and drug status). The Benjamini-Hochberg method (Benjamini and Hochberg, 1995) was employed to mitigate the false discovery rate (FDR) across several tests. We performed a binary logistic regression analysis to examine the predictors of deficit schizophrenia (dependent variable) against controls, presenting odds ratios with 95% confidence intervals. The analyses were bootstrapped (n=5000), and the bootstrapped results are reported where discrepancies between the latter and the classical approaches exist. All analyses were two-tailed and a significance level of p=0.05 was considered significant. The primary analysis was the multiple regression analysis of OSOS on the 5 BI biomarkers allowing for the effects of possible confounding variables. The power analysis, performed utilizing G*Power version 3.1.9.7, reveals that a minimum sample size of 103 participants is required for conducting a multiple regression analysis that includes three predictors, an effect size of 0.111 (explains around 10% of the variance), an alpha level of 0.05, and a desired power of 0.8.

## Results

### Socio-demographic data, neuropsychiatric scales, and serum TRYCATs

The demographic and clinical data in healthy controls (HC), patients with MNP and SNP are presented in **Table 1**. The results show no significant difference in age, BMI, sex ratio, smoking, and residency among the study groups. The patient group had significantly lower education years and married subjects than the controls. Psychosis, hostility, excitement, mannerism, the PANSS negative domain score, total SANS, and OSOS differed among the study groups with the highest values in MNP, followed by SNP, and the lowest values in the healthy control group. TRP and 3-HK concentrations were significantly different between the three study groups with both levels decreasing from control to SNP and MNP. Both MNP and SNP patients experienced a significant decrease in KYN compared with the control group. Serum KA was significantly decreased in the MNP group compared with the control group. The MNP group shows a significant increase in AA and QA, and a decrease in 3-HAA compared with the SNP and healthy control groups.

**Table 1.** Demographic and clinical data in healthy controls (HC), patients with major neuro-cognitive psychosis (MNP), and simple neuro-cognitive psychosis (SNP).

| Variables | Controls<br>N=60 | SNP<br>N=68 | MNP<br>N=52 | F | df | p |
| --- | --- | --- | --- | --- | --- | --- |
| Age (years) | 38.6±10.9 | 41.2±10.9 | 41.2±8.6 | 1.30 | 2/177 | 0.276 |
| BMI (kg/m<sup>2</sup>) | 27.36±4.70 | 27.74±4.27 | 27.81±3.08 | 0.20 | 2/177 | 0.818 |
| Sex F/M | 18/42 | 22/46 | 10/42 | 2.75 | 2 | 0.253 |
| Education (yrs). | 10.9±3.7<sup>B,C</sup> | 7.7±3.2<sup>A</sup> | 7.3±3.7<sup>A</sup> | 18.80 | 2/177 | <0.001 |
| Smoking No/Yes | 41/19 | 46/22 | 31/21 | 1.15 | 2 | 0.563 |
| Single/Married | 11/49 | 39/29 | 29/23 | 23.90 | 2 | <0.001 |
| Rural/Urban | 5/55 | 16/52 | 9/43 | 5.32 | 2 | 0.070 |
| Psychosis | -6.625±0.060<sup>B,C</sup> | 1.120±2.176<sup>A,C</sup> | 6.179±1.987<sup>A,B</sup> | 802.38 | 2/177 | <0.001 |
| Hostility | -5.364±0.276<sup>B,C</sup> | 1.351±2.457<sup>A,C</sup> | 4.423±2.233<sup>A,B</sup> | 382.73 | 2/177 | <0.001 |
| Excitement | -4.083±0.399<sup>B,C</sup> | 0.701±1.847<sup>A,C</sup> | 3.795±1.984<sup>A,B</sup> | 359.63 | 2/177 | <0.001 |
| Mannerism | -2.08±0<sup>B,C</sup> | 0.533±1.431<sup>A,C</sup> | 1.703±1.684<sup>A,B</sup> | 134.94 | 2/177 | <0.001 |
| PANSS negative | 0.450±0.594<sup>B,C</sup> | 19.074±4.198<sup>A,C</sup> | 30.308±4.913<sup>A,B</sup> | 938.39 | 2/177 | <0.001 |
| Total SANS | 6.1±4.3<sup>B,C</sup> | 65.6±9.5<sup>A,C</sup> | 84.9±7.0<sup>A,B</sup> | 1805.88 | 2/177 | <0.001 |
| OSOS | -1.292±0.037<sup>B,C</sup> | 0.296±0.307<sup>A,C</sup> | 1.104±0.248<sup>A,B</sup> | 1574.60 | 2/177 | <0.001 |
| Tryptophan (z scores) | 0.455±1.153<sup>B,C</sup> | -0.031±0.821<sup>A,C</sup> | -0.485±0.775<sup>A,B</sup> | 14.18 | 2/177 | <0.001 |
| Kynurenine (z scores) | 0.343±0.900<sup>B,C</sup> | -0.028±1.069<sup>A</sup> | -0.358±0.894<sup>A</sup> | 7.38 | 2/177 | 0.001 |
| 3-hydroxy-kynurenine (z scores) | 0.381±0.804<sup>B,C</sup> | -0.029±1.035<sup>A,C</sup> | -0.401±1.009<sup>A,B</sup> | 9.37 | 2/177 | <0.001 |
| Kynurenic acid (z scores) | 0.359±1.179<sup>C</sup> | 0.151±0.969 | -0.612±0.323<sup>A</sup> | 16.96 | 2/177 | <0.001 |
| Anthranilic acid (z scores) | -0.221±0.656<sup>C</sup> | -0.052±0.960<sup>C</sup> | 0.322±1.278<sup>A,B</sup> | 4.41 | 2/177 | 0.013 |
| 3-hydroxy-anthranilic acid (z scores) | 0.186±0.826<sup>C</sup> | 0.181±1.093<sup>C</sup> | -0.451±0.927<sup>A,B</sup> | 8.04 | 2/177 | <0.001 |
| Quinolinic acid (z scores) | -0.332±0.592<sup>C</sup> | -0.123±1.068<sup>C</sup> | 0.544±1.074<sup>A,B</sup> | 13.07 | 2/177 | <0.001 |

All values are shown as mean (SD). All quantitative data are expressed in z scores, except age, body mass index (BMI).
<sup>A,B,C</sup> : post-hoc comparisons among group means.
PANSS: The Positive and Negative Syndrome Scale, SANS: The Scale for the Assessment of Negative Symptoms, OSOS: overall severity of schizophrenia.

### Brain injury biomarkers in MNP and SNP

**Table 2** shows the comparison of serum BI markers in the three study groups. NSE, GFAP, Ptau217, and the ND index were significantly different between the three study groups and increased from controls to SNP to MNP. S100B was significantly higher in MNP than in controls and SNP. There were no significant differences in nestin levels between the three study groups. **Electronic Supplementary File, Table 1** shows increased serum levels of pTau217, NSE, GFAP and ND index in SNP+MNP (schizophrenia) as compared with controls. There were no significant differences in S100B and nestin between the study groups.

**Table 2.** Associations between brain injury biomarkers and three diagnostic groups, namely simple neuro-cognitive psychosis (SNP), major neuro-cognitive psychosis (MNP), and healthy controls (HC).

| Parameter | Controls<br>n=60 | SNP<br>n=68 | MNP<br>n=52 | F | df | p |
| --- | --- | --- | --- | --- | --- | --- |
| S100B (pg/ml) | 14.89 (2.54)<sup>C</sup> | 13.83 (2.38)<sup>C</sup> | 28.32 (2.73)<sup>A,B</sup> | 3.45 | 2/177 | 0.034 |
| pTau217 (pg/ml) | 3.10 (0.19)<sup>C</sup> | 3.27 (0.18)<sup>C</sup> | 4.51 (0.20)<sup>A,B</sup> | 12.40 | 2/177 | <0.001 |
| Nestin (pg/ml) | 71.91 (5.30) | 67.91 (4.96) | 72.86 (5.70) | 0.26 | 2/177 | 0.774 |
| NSE (pg/mL) | 0.48 (0.03)<sup>B,C</sup> | 0.73 (0.03)<sup>A,C</sup> | 0.95 (0.04)<sup>A,B</sup> | 46.99 | 2/177 | <0.001 |
| GFAP (pg/ml) | 1.30 (0.48)<sup>B,C</sup> | 2.72 (0.45)<sup>A,C</sup> | 6.03 (0.52)<sup>A,B</sup> | 62.51 | 2/177 | <0.001 |
| BII (z score) | -1.763 (0.267)<sup>B,C</sup> | -0.298 (0.25)<sup>A,C</sup> | 2.425 (0.287)<sup>A,B</sup> | 57.70 | 2/177 | <0.001 |

All data are shown as estimated marginal mean (SE). All results of GLM analysis (adjusted for sex, age, smoking and BMI)
<sup>A,B,C</sup> : post-hoc comparisons among group means.
GFAP: Glial fibrillary acidic protein, S100B: S100 calcium-binding protein B, NSE: Neuron-Specific Enolase, pTau217: phosphorylated tau protein at amino acid 217. BII: brain injury index

To delineate the best predictors of MNP+SNP (schizophrenia) versus controls and MNP versus SNP, we have performed binary logistic regression analyses with schizophrenia and MNP as the dependent variables (and controls or SNP, respectively, as reference groups) using an automatic stepwise method with the damage biomarkers with and without TRYCATs as explanatory variables, while allowing for the effects of age, sex, BMI, and TUD (**Table 3**). Regression #1 shows that schizophrenia was best predicted by increased NSE and GFAP (χ^2^=99.511, df=2, p<0.001) with a Nagelkerke effect size of 0.590, and overall accuracy of 82.2%. Regression #2 shows that MNP was best differentiated from SNP by increased levels of pTau217, NSE, and GFAP (χ^2^ =52.542, df=3, p<0.001, Nagelkerke R^2^=0.476) with an overall accuracy 76.7%. Increases in NSE and GFAP coupled with decreases in TRP and 3HK predict MNP+SNP versus controls (χ^2^ =112.609, df=4, p<0.001, Nagelkerke R^2^=0.646) with an overall accuracy of 86.7%, sensitivity of 90% and specificity of 80% (Regression #3). MNP (reference group SNP) is significantly predicted by increases in pTau217 and NSE and lowered levels of TRP and KA (Regression #4). The overall accuracy of the prediction of MNP according is 82.5% with a sensitivity of 80.8% and a specificity of 83.8% (χ^2^ =72.900, df=4, p<0.001, Nagelkerke R^2^=0.611).

**Table 3.**
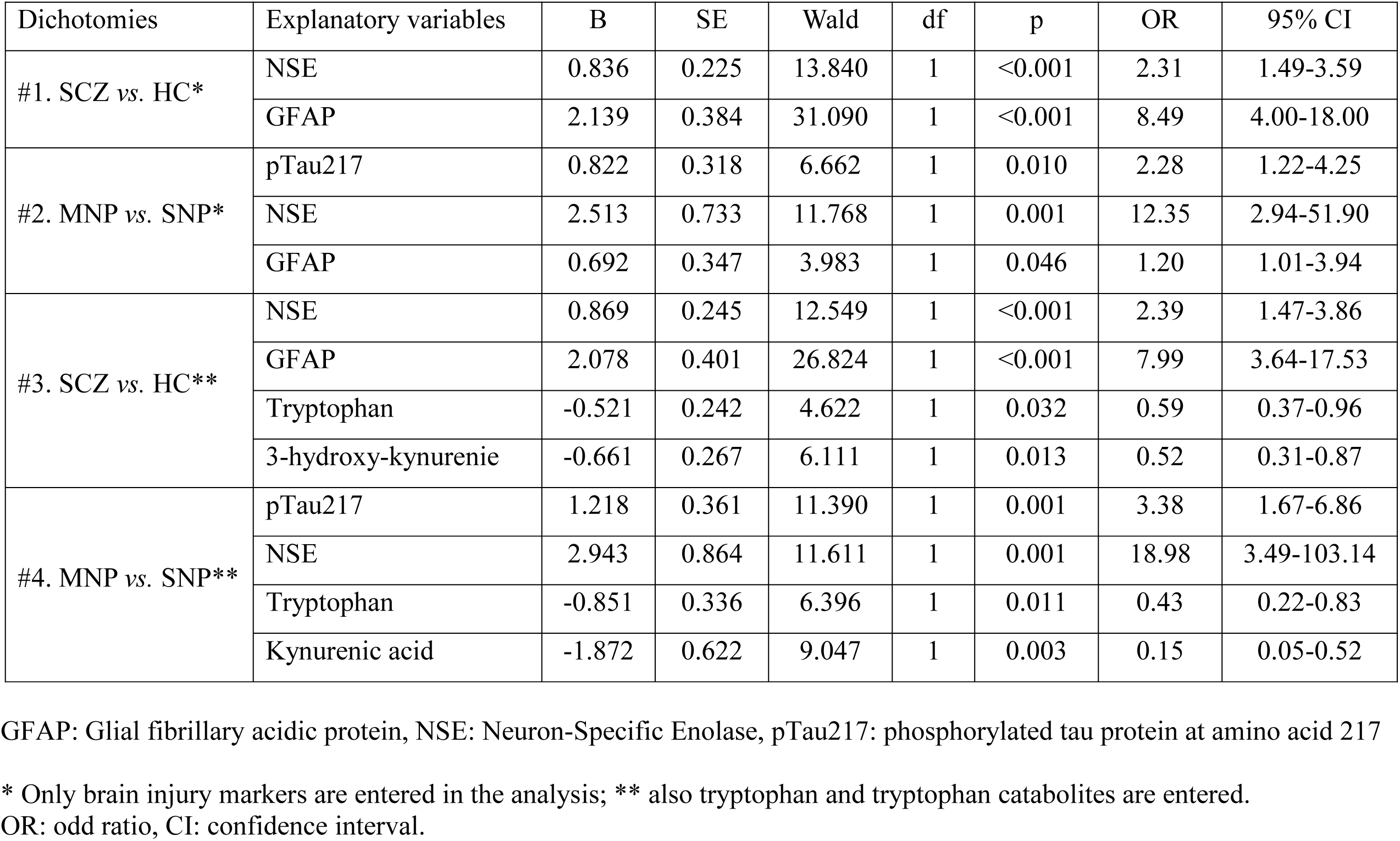
Results of binary logistic regression analyses with schizophrenia (SCZ) or its subtypes (major neuro-cognitive psychosis (MNP), and simple neuro-cognitive psychosis (SNP)) as a dependent variable and healthy controls (HC) as reference group

We have also examined whether the drug status of the patients may have affected the outcome of the BI biomarkers. We could not find any effects of clozapine (n=59, F=0.76, df=5/109, p=0.578), olanzepine (n=73, F=0.45, df=5/109, p=0.810), flufenazine (n=27, F=1.05, df=5/109, p=0.391), risperidal (n=27, F=0.68, df=5/109, p=0.640), haloperidol (n=28, F=1.59, df=5/108, p=0.170), trifluoperazine (n=29, F=0.29, df=5/106, p=0.917), carbamazepine (n=45, F=0.50, df=5/109, p=0.778), escitalopram (n=33, F=0.09, df=5/108, p=0.994) and propranolol (n=56, F=0.96, df=5/109, p=0.449) (all results of multivariate GLM analysis conducted in MNP+SNP patients). Moreover, not one of the tests of between-subject effects was significant even without FDR p correction.

### Prediction of OSOS by brain damage markers

**Table 4** shows different stepwise multiple regression analyses with OSOS as dependent variable, and serum BI markers, TRP and TRYCATs as explanatory variables while allowing for the effects of age, sex, BMI, and TUD. Regression #1 shows that 54.2% of the variance in OSOS in all subjects was explained by the regression on GFAP and NSE. Regression #2 shows that 25.6% of the variance in the OSOS in MNP+SNP can be explained by NSE and the ND index. **Figure 1 and Figure 2** show the partial regressions of the OSOS score on NSE and GFAP, respectively. Regression #3 shows that 58.4% of the variance in OSOS in all subjects was explained by GFAP, NSE (positively), TRP, and 3-HK (inversely). NSE, S100B (both positively), KA and TRP (both negatively associated), explained together 35.4% of the variance in OSOS in schizophrenia patients (Regression #4).

**Figure 1.**
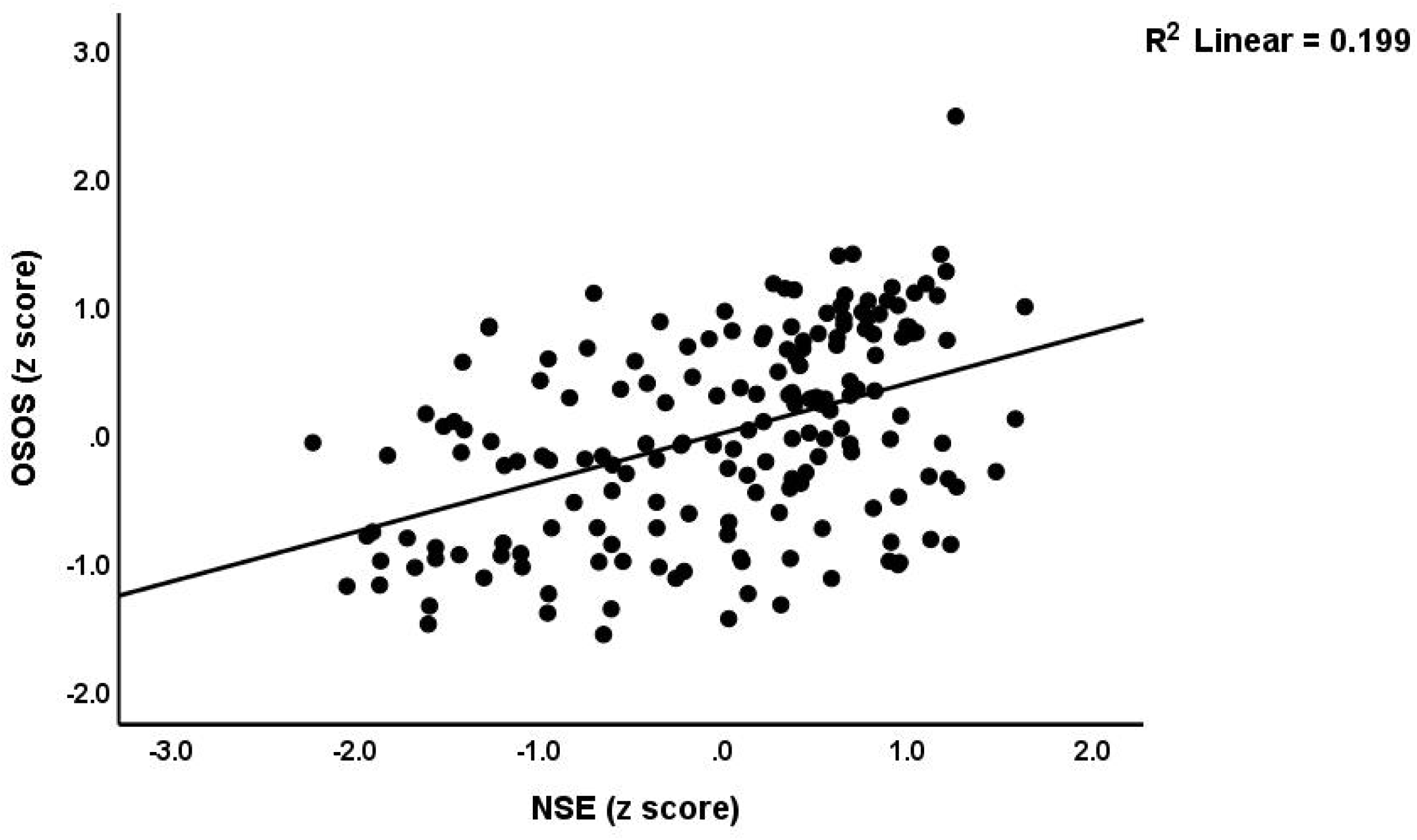
Partial regression of the overall severity of schizophrenia (OSOS) score on neuron-specific

**Figure 2.**
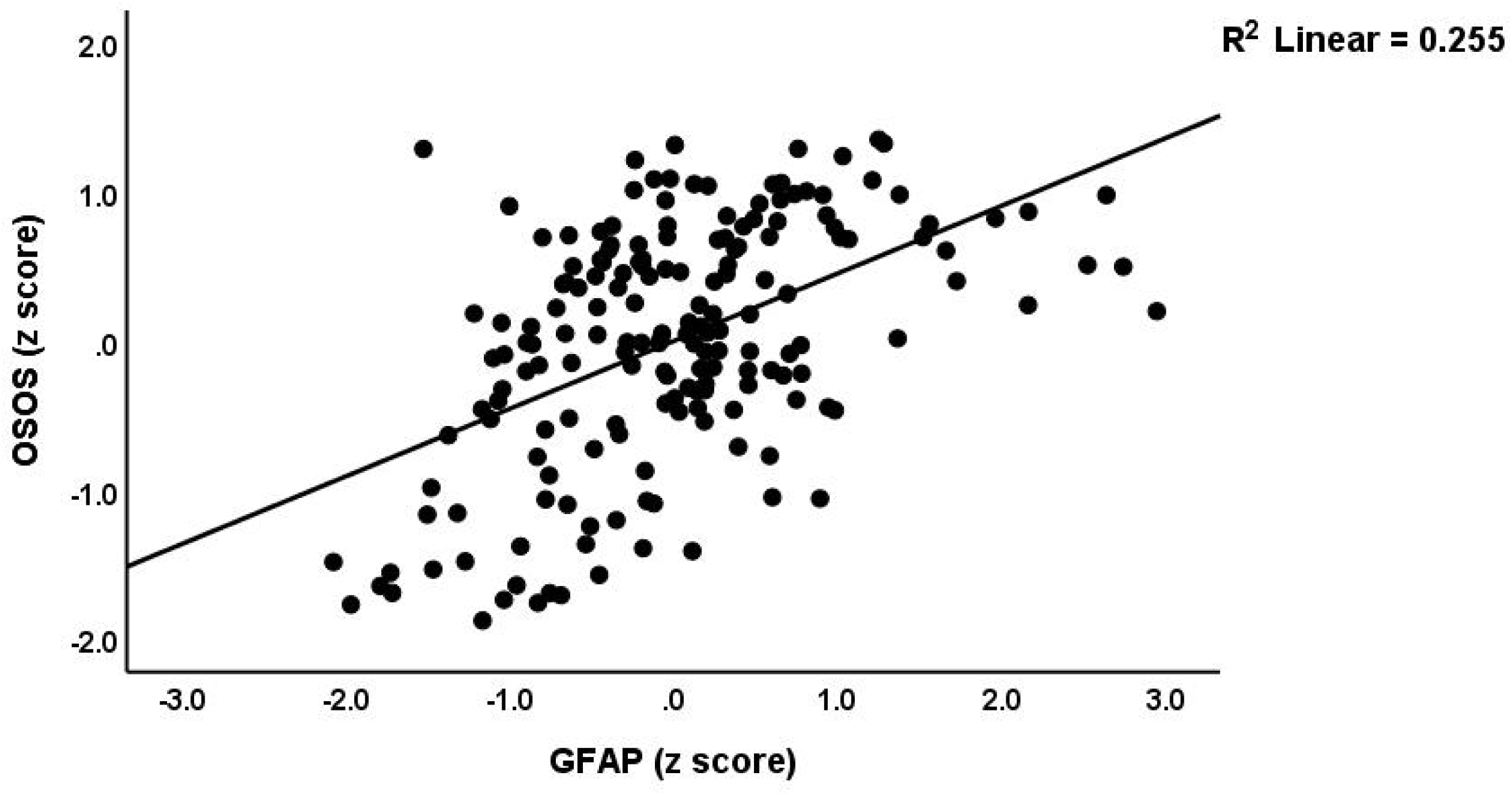
Partial regression of the overall severity of schizophrenia (OSOS) score on glial fibrillary

**Table 4.** Results of multiple regression with the overall severity of schizophrenia (OSOS) as the dependent variable, and serum brain injury biomarkers, tryptophan, and tryptophan catabolites as explanatory variables.

| Dependent Variables | Explanatory Variables | $\beta$ | t | p | F model | df | p | R<sup>2</sup> |
| --- | --- | --- | --- | --- | --- | --- | --- | --- |
| #1. OSOS in all subjects | **Model** |  |  |  | 104.85 | 2/177 | <0.001 | 0.542 |
|  | GFAP | 0.499 | 8.85 | <0.001 |  |  |  |  |
|  | NSE | 0.368 | 6.54 | <0.001 |  |  |  |  |
| #2. OSOS in MNP+SNP | **Model** |  |  |  | 20.13 | 2/117 | <0.001 | 0.256 |
|  | NSE | 0.307 | 3.16 | 0.002 |  |  |  |  |
|  | BII | 0.264 | 2.72 | 0.007 |  |  |  |  |
| #3. OSOS in all subjects | **Model** |  |  |  | 61.30 | 4/175 | <0.001 | 0.584 |
|  | GFAP | 0.443 | 7.96 | <0.001 |  |  |  |  |
|  | NSE | 0.350 | 6.45 | <0.001 |  |  |  |  |
|  | Tryptophane | -0.166 | -3.28 | 0.001 |  |  |  |  |
|  | 3-hydroxy-kynurenine | -0.111 | -2.23 | 0.027 |  |  |  |  |
| #4. OSOS in MNP+SNP | **Model** |  |  |  | 15.79 | 4/115 | <0.001 | 0.354 |
|  | NSE | 0.341 | 4.241 | <0.001 |  |  |  |  |
|  | Kynurenic acid | -0.268 | -3.360 | 0.001 |  |  |  |  |
|  | S100B | 0.189 | 2.518 | 0.013 |  |  |  |  |
|  | Tryptophane | -0.187 | -2.449 | 0.016 |  |  |  |  |

GFAP: Glial fibrillary acidic protein, NSE: Neuron-Specific Enolase, pTau217: phosphorylated tau protein at amino acid 217; BII: brain imaging index.

### Intercorrelation matrix among damage markers and TRYCATs

The intercorrelation matrix of the correlation coefficients of the BI markers and TRYCATs is presented in **Table 5**. The results show that QA and zQA-zKA are significantly correlated with pTau217, NSE, GFAP, and BD index. Serum 3-HK was significantly and inversely correlated with NSE, GFAP, and the BD index. TRP, KYN, and KA were significantly and inversely correlated with NSE, GFAP, and the BD index. Serum 3-HK was significantly and inversely correlated with GFAP and the BD index. AA did not show any significant correlations with any measured BD markers.

**Table 5.** Intercorrelations among brain injury biomarkers and tryptophan catabolites.

| Variables | **pTau217** | **NSE** | **GFAP** | **BII** |
| --- | --- | --- | --- | --- |
| Tryptophan | 0.038 (0.61) | **-0.206 (0.006)** | **-0.245 (0.001)** | **-0.176 (0.018)** |
| Kynurenine | -0.071 (0.342) | **-0.339 (<0.001)** | **-0.269 (<0.001)** | **-0.218 (0.003)** |
| 3-hydroxy-kynurenine | **-0.155 (0.038)** | -0.128 (0.087) | **-0.158 (0.034)** | **-0.173 (0.020)** |
| Kynurenine Acid (KA) | -0.117 (0.118) | **-0.223 (0.003)** | **-0.226 (0.002)** | **-0.259 (<0.001)** |
| Anthranilic acid | -0.014 (0.848) | 0.132 (0.078) | 0.112 (0.135) | 0.12 (0.109) |
| 3-hydroxy-anthranilic acid | -0.126 (0.093) | -0.145 (0.053) | **-0.230 (0.002)** | **-0.205 (0.006)** |
| Quinolinic acid (QA) | **0.160 (0.031)** | **0.261 (<0.001)** | **0.276 (<0.001)** | **0.262 (<0.001)** |
| zQA-zKA | **0.234 (0.002)** | **0.346 (<0.001)** | **0.368 (<0.001)** | **0.377 (<0.001)** |

GFAP: Glial fibrillary acidic protein, NSE: Neuron-Specific Enolase, pTau217: phosphorylated tau protein at amino acid 217; BII: brain injury index; zQA-zKA: index for the QA / KA ratio.

### Prediction of brain damage markers by TRP and TRYCATs

**Table 6** shows the results of multiple regression with the brain damage markers as dependent variables, and serum TRP and TRYCATs as explanatory variables. Regression #1 shows that 6.1% of the variance in the pTau217 was explained by the regression on sex and the composite score zQA–z KA. Regressions #2 shows that 21.2% of the variance in NSE can be explained by zQA–zKA and KYN. Regression #3 shows that 22.4% of the variance in the GFAP was explained by zQA–z KA, KYN, and TRP. **Figure 3** shows the partial regression of GFAP on zQA–zKA.

**Figure 3.**
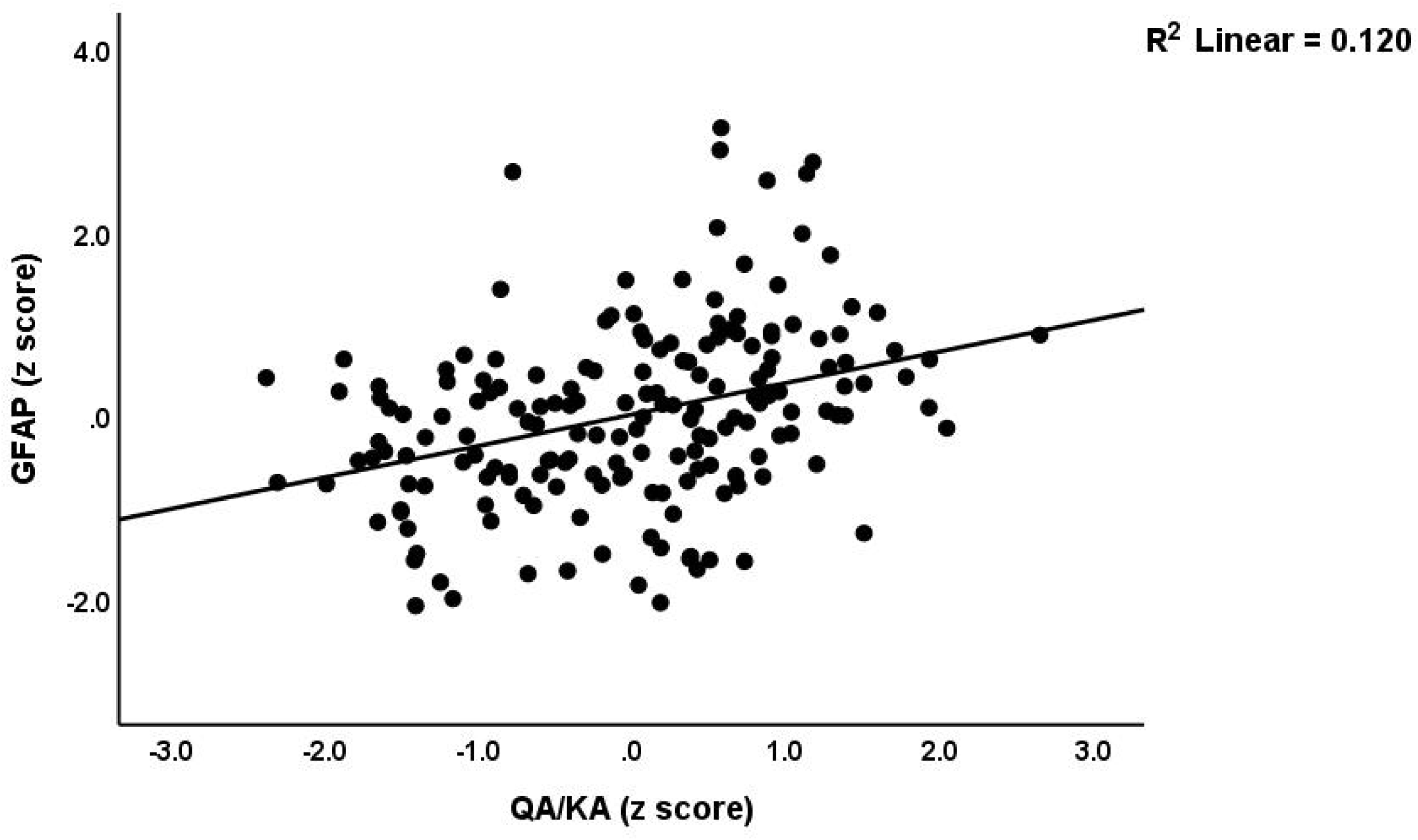
Partial regression of serum glial fibrillary acidic protein (GFAP) levels on an index of the ratio quinolinic acid (QA) on kynurenic acid (KA) (adjusted for age, sex, smoking and BMI), p<0.001.

**Table 6.**
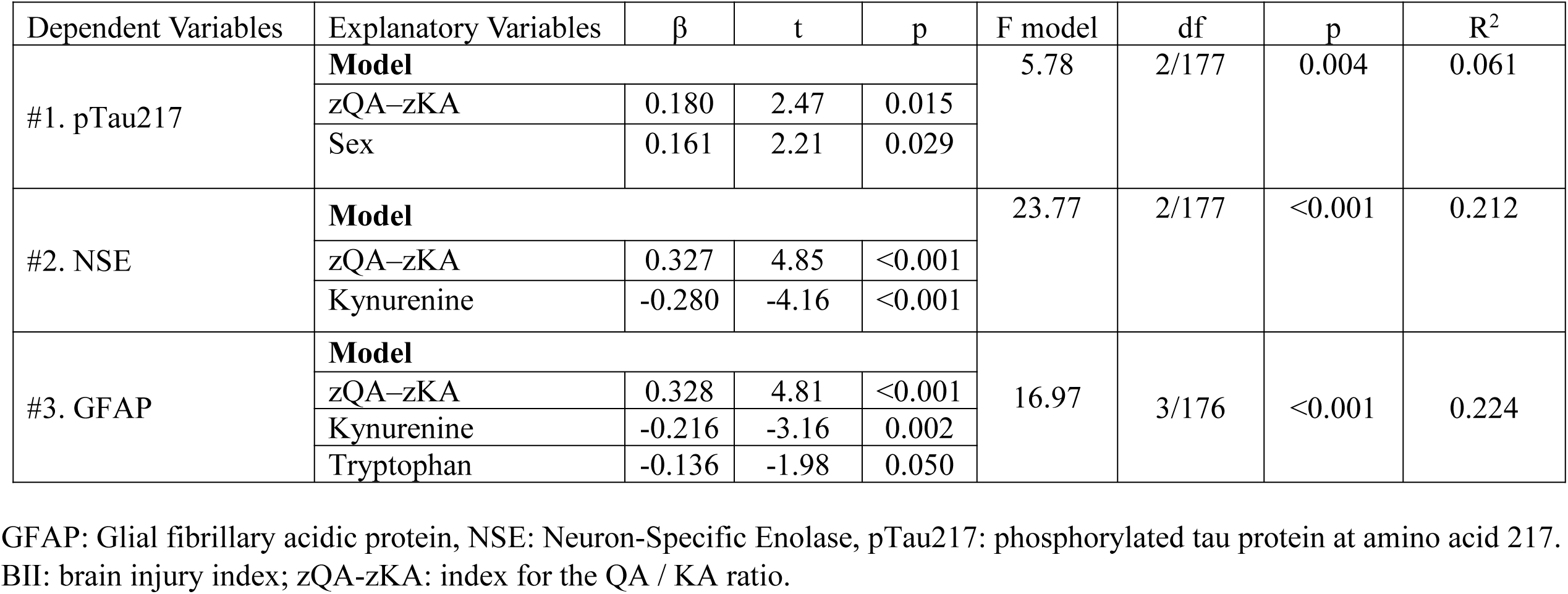
Results of multiple regression with brain injury biomarkers as dependent variables, and serum tryptophan and tryptophan catabolites as explanatory variables.

## Discussion

### Brain injury biomarkers in MNP, SNP and controls

The first significant discovery of the current investigation is that the serum levels of brain injury markers were elevated in MNP and SNP in comparison to controls. To be more precise, the SNP+MNP (schizophrenia) group exhibited substantially higher serum levels of pTau217, NSE, and GFAP in comparison to healthy controls, whereas no difference in serum S100B and nestin between the groups were observed. However, the sample was further divided into patients with MNP and SNP, which revealed numerous additional significant differences. As a result, the BII, NSE, and GFAP were substantially higher in MNP than in SNP, and in both MNP or SNP than in controls. In addition, the serum S100B levels in MNP were substantially higher than those in controls and SNP. Phrased differently, if we had not separated MNP and SNP into distinct categories we would have missed the most important information. These findings further validate the differentiation between the two distinct subclasses of neurocognitive psychoses.

It can be deduced that MNP demonstrates much larger brain astroglial injuries (GFAP and S100B), axonal injuries (nestin, an intermediate filament protein), and neuronal damage (NSE, an enzyme secreted during neuronal injuries) compared to SNP. To accurately ascertain the differential functions of differentially expressed proteins (DEPs), we conducted protein-protein interaction (PPI) network analysis, thereafter, performing enrichment and annotation analysis utilizing String (https://cn.string-db.org/). This PPI network analysis (minimum required interaction score of 0.4) performed on the four DEPs discriminating MNP from SNP, shows that a tight cluster could be formed (average local clustering coefficient of 1, expected number of edges=0, number of edges=6, PPI enrichment p=8.38e-6). Functional enrichment analysis showed that this network of 4 genes was strongly associated with cell body (p=0.0015, all 4 DEPs included) and glial cell projections (p=0.0246, only MAPT and GFAP are included). A second functional enrichment analysis performed on these 4 DEPs and 5 interacting DEPs in the first shell showed again a tight network, with an average local clustering coefficient of 0.952, and average node degree of 7.56 (expected number of edges = 6, edges=34, enrichment p=2.24e-14). Regulation of synaptic plasticity (p=0.0045; GFAP, S100B and MAPT or pTau217), regulation of neurogenesis (p=0.0176, GFAP and MAPT are included) and distal axons (p=0.0015, ENO2 and MAPT) are significantly enriched in this gene network. On the other hand, enrichment analysis performed on the two genes that separate SNP+MNP from controls (namely NSE and GFAP) showed a strong association with type III intermediate neurofilament (p=0.00078). In general, these functional analyses demonstrate that MNP is associated with a greater degree of damage to cell bodies, axons, glial cell projections, and reduced neurogenesis and synaptic plasticity in comparison to SNP and controls. Additionally, SNP patients exhibit significantly more damage to neuronal and astroglial projections than controls. Therefore, it seems that neuronal cell injury is present in both phenotypes; however, the damage in MNP may be more extensive and affects other cell structures.

It was previously observed that GFAP levels are elevated in schizophrenia patients relative to controls and are associated with psychotic symptoms (Feresten et al., 2013). Moreover, serum NSE concentrations are elevated in patients with treatment-resistant schizophrenia compared to healthy controls (Medina-Hernández et al., 2007, Liu et al., 2020). Phosphorylated tau protein has been observed to be elevated in the cerebrospinal fluid of schizophrenic patients (Schönknecht et al., 2003), although another study reported a reduction in total and pTau217 levels in these individuals (Demirel Ö et al., 2017). There are also some negative results showing no significant difference between schizophrenia patients and controls in the serum and CSF levels of GFAP and NSE (Steiner et al., 2006). The latter results may not be statistically solid due to the lower number of patients included in the study (n=12 only). Nevertheless, any discrepancies in results between schizophrenia patients and controls in those studies may be attributed to the failure to categorize the MNP+SNP study group into its relevant separate classes.

### Associations among brain injury biomarkers and the TRYCAT pathway

The second major finding of this study is that increased QA levels and lowered KA, KYN or 3HAA levels predict brain injury markers such as pTau217, NSE and GFAP. Nevertheless, the best predictor of these three brain injury markers is the ratio QA/KA. In theory and as reviewed in the Intro, an elevation in QA and a reduction in KA may induce NMDAR activation, subsequently promoting the production of reactive oxygen species under mitochondrial oxidative stress, and resulting in cellular damage and brain injury (Yu et al., 2016, Cao et al., 2021, Kindler et al., 2020). Nevertheless, not all TRYCATS, including QA, can pass an intact BBB, although schizophrenia is accompanied by a breakdown of the BBB (Maes et al., 2019b). The latter may allow QA and other TRYCATs to pass the BBB more quickly suggesting that the peripheral increases in QA versus KA may have an effect including at the NMDARs. Such responses may induce excitotoxity and neurotoxicity leading to brain cell injuries in neurons and astroglia.

### IRS activation, oxidative stress, and TRYCATs in MNP

It is important to emphasize that both QA and the activation of IRS, along with heightened oxidative stress, significantly contribute to enhanced neurotoxicity and the phenome of MNP (this study, Roomruanwong et al., 2018; Popov et al., 2024). The inflammatory response can induce activation of microglia and astrocytes, leading to the release of various pro-inflammatory mediators that may contribute to neuronal dysfunction and the advancement of CNS pathology (Khansari et al., 2009). Activated glial cells secrete several pro-inflammatory mediators that may lead to neuronal dysfunction and injury (Khansari et al., 2009). It is plausible to conjecture that in MNP, augmented QA, diminished KA, IRS activation, and heightened oxidative stress led to neuronal and astroglial injuries. In major depression, the impact of heightened peripheral inflammation on the physio-affective phenome is mediated by biomarkers of brain damage, including raised plasma levels of neurofilament light chain (NFL), GFAP, and P-tau217 (Al-Hakeim et al., 2023). Additionally, several brain damage markers, notably S100B, are implicated in the immune system and the inflammatory etiopathogenesis of schizophrenia (Rothermundt et al., 2009; Steiner et al., 2009; Mondelli and Howes, 2014; Monji et al., 2009). Furthermore, some pro-inflammatory cytokines may increase S100B secretion (de Souza et al., 2013). Furthermore, oxidative stress exerts detrimental effects on astroglial and neuronal cells, as well as influencing the synthesis and secretion of S100B (Masmoudi-Kouki et al., 2011). S100B is predominantly expressed in astrocytes, which serve as a significant source of free radicals in the central nervous system (CNS) (Lohr, 1991). Consequently, high S100B may either contribute to oxidative stress or serve as a compensatory mechanism in response to heightened oxidative stress (District, 2019).

Additionally, as discussed in the Introduction, the TRYCAT pathway is primarily a protective pathway that safeguards against oxidative stress and inflammation. MNP patients are more susceptible to the adverse effects of inflammatory mediators, oxidative stress, and QA due to the reduced levels of TRP (which protects against microbiota infections), KYN, KA, and 3HK (which possess anti-inflammatory and antioxidant properties respectively). As recently reported in a meta-analysis, the TRYCATs pathway may be activated in certain patients with MNP+SNP (Almulla et al., 2022). In MNP+SNP, a low-grade immune response seems to be associated with a dysbalance in the activation of the IDO and the TRYCATs pathway resulting in increased production of KA in schizophrenia (Muller and J Schwarz, 2010, Anderson and Maes, 2013). Conversely, IgA responses to neurotoxic TRYCATs are markedly elevated in MNP compared to SNP and correlate with negative and PHEM (psychosis, hostility, excitation, and mannerism) symptoms, as well as affective (depression and anxiety) symptoms, episodic memory deficits, and the generation of false memories (Kanchanatawan et al., 2018c).

### Limitations

The findings of the present investigation, conducted in Iraq, merit replication among MNP patients compared to SNP patients and controls in other countries and cultures. The findings presented here would have been even more interesting if we had examined indicators of M1 macrophage and T helper 1 cytokines, coupled with LPS of some Gram-negative bacteria, such as *Klebsiella pneumoniae*, in peripheral blood (Maes et al., 2021). Activated M1 and T helper1 cells may produce neurotoxic cytokines that activate the TRYCAT pathway, whilst LPS in itself is neurotoxic and may activate the TRYCAT pathway (Maes et al., 2021).

## Conclusions

The findings of the current study further substantiate the distinction between the two distinct schizophrenia subclasses, namely major neurocognitive psychosis (MNP) versus simple neurocognitive psychosis). Studies that do not differentiate between MNP and SNP and lump both groups together are difficult to interpret. Our findings indicate that levels of NSE and GFAP were markedly elevated in MNP compared to SNP, and in both MNP and SNP relative to the control group. The levels of Serum S100B were significantly elevated in MNP when compared to both the control group and SNP. The elevation of QA levels alongside a reduction in KA predicts a rise in pTau217, NSE, and GFAP levels. The ratio of QA to KA serves as the most reliable predictor among these three markers of brain injury. This suggests that elevation of peripheral QA levels may play a role in the etiology of brain injuries observed in MNP. These results show that damage to neurofilaments is a new drug target to treat MNP and SNP and that injuries to neuron bodies, neuronal and astroglial projections, aberrations in neuroplasticity and neurogenesis, and increase QA levels are new drug targets to treat MNP.

## Supporting information

ESF, Table 1. ESF, Table 2.

## Data Availability

The data set produced and/or examined in the present work will be accessible from the corresponding author (M.M) upon reasonable request, after the authors' complete utilization of the dataset.

## Acknowledgements

We express our gratitude to the staff at Ibn-Rushd Hospital for their support throughout the sample collection procedure. Furthermore, we extend our gratitude to the proficient personnel at the Asia Clinical Laboratory in Najaf City for their support with the ELISA measurements.

## Ethics approval and consent to participate

The research conducted in this study has been approved by the ethics committee (IRB) of the College of Science, University of Kufa, Iraq (T4220/2023). This approval is in accordance with the International Guideline for Human Research Protection, as mandated by the Declaration of Helsinki. Before participating in this study, each subject provided written informed consent.

## Consent to publish

The authors assert that human study participants granted informed permission for the publishing of their anonymized findings.

## Author’s contributions

Hussein Kadhem Al-Hakeim: conceptualization; formal analysis; writing - original draft. Ameer Abdul Razzaq Al-Issa: conceptualization, data curation. Mengqi Niu: writing - review & editing. Yingqian Zhang: writing - review & editing. Michael Maes: conceptualization; supervision; writing - review & editing. All authors approved the submitted manuscript.

## References

Al-Hakeim, H. K., Al-Naqeeb, T. H., Almulla, A. F. & Maes, M. J. J. O. A. D.2023. The physio-affective phenome of major depression is strongly associated with biomarkers of astroglial and neuronal projection toxicity which in turn are associated with peripheral inflammation, insulin resistance and lowered calcium. 331, 300–312.

Al-Hakeim, H. K. A.-I., A. A.; Chen, C.; MAES M 2024. The tryptophan catabolite pathway in major and simple neurocognitive psychosis: a double-edged sword with two sharpened edges. Research Gate, DOI: 10.13140/RG.2.2.16389.15841.

Almulla, A. F., Al-Hakeim, H. K. & Maes, M. 2021. Schizophrenia phenomenology revisited: positive and negative symptoms are strongly related reflective manifestations of an underlying single trait indicating overall severity of schizophrenia. CNS spectrums,26, 368–377.

Almulla, A. F., Vasupanrajit, A., Tunvirachaisakul, C., Al-Hakeim, H. K., Solmi, M., Verkerk, R. & Maes, M. 2022. The tryptophan catabolite or kynurenine pathway in schizophrenia: meta-analysis reveals dissociations between central, serum, and plasma compartments. Mol Psychiatry, 27, 3679–3691.

Anderson, G. & Maes, M. 2013. Schizophrenia: linking prenatal infection to cytokines, the tryptophan catabolite (TRYCAT) pathway, NMDA receptor hypofunction, neurodevelopment and neuroprogression. Progress in Neuro-Psychopharmacology and Biological Psychiatry, 42, 5–19.

Andreasen, N. C. 1989. The Scale for the Assessment of Negative Symptoms (SANS): conceptual and theoretical foundations. The British Journal of Psychiatry,155, 49–52.

Benjamini, Y. & Hochberg, Y. 1995. Controlling the false discovery rate: a practical and powerful approach to multiple testing. Journal of the Royal statistical society: series B (Methodological), 57, 289–300.

Bornstein, S. R., Berger, I. & Steenblock, C. J. S. 2020. Are Nestin-positive cells responsive to stress? 23, 662–666.

Brenner, M. 2014. Role of GFAP in CNS injuries. Neuro science letters, 565, 7–13.

Cao, B., Chen, Y., Ren, Z., Pan, Z., Mcintyre, R. S. & Wang, D. 2021. Dysregulation of kynurenine pathway and potential dynamic changes of kynurenine in schizophrenia: A systematic review and meta-analysis. Neuro science & Biobehavioral Reviews,123, 203–214.

Das Neves, S. P., Sousa, J. C., Sousa, N., Cerqueira, J. J. & Marques, F. J. G. 2021. Altered astrocytic function in experimental neuroinflammation and multiple sclerosis. 69, 1341–1368.

De Souza, D. F., Wartchow, K., Hansen, F., Lunardi, P., Guerra, M. C., Nardin, P. & Gonçalves, C.-A. 2013. Interleukin-6-induced S100B secretion is inhibited by haloperidol and risperidone. Progress in Neuro-Psychopharmacology and Biological Psychiatry, 43, 14–22.

Demirel Ö, F., Cetin, I., Turan, Ş., Yildiz, N., Sağlam, T. & Duran, A. 2017. Total Tau and Phosphorylated Tau Protein Serum Levels in Patients with Schizophrenia Compared with Controls. Psychiatr Q, 88, 921–928.

District, C. 2019. Serum levels of oxidants and protein S100B were associated in lGlClthe first-episode drug naïve patients with schizophrenia Global Clin. Translational Res, 84–92.

Feresten, A. H., Barakauskas, V., Ypsilanti, A., Barr, A. M. & Beasley, C. L. 2013. Increased expression of glial fibrillary acidic protein in prefrontal cortex in psychotic illness. Schizophrenia research,150, 252–257.

González-Cota, A. L., Martínez-Flores, D., Rosendo-Pineda, M. J. & Vaca, L. 2024. NMDA receptor-mediated Ca2+ signaling: Impact on cell cycle regulation and the development of neurodegenerative diseases and cancer. Cell Calcium, 102856.

Huibregtse, M. E., Bazarian, J. J., Shultz, S. R. & Kawata, K. 2021. The biological significance and clinical utility of emerging blood biomarkers for traumatic brain injury. Neuroscience & Biobehavioral Reviews, 130, 433–447.

Jiang, Y., Luo, C., Wang, J., Palaniyappan, L., Chang, X., Xiang, S., Zhang, J., Duan, M., Huang, H., Gaser, C., Nemoto, K., Miura, K., Hashimoto, R., Westlye, L. T., Richard, G., Fernandez-Cabello, S., Parker, N., Andreassen, O. A., Kircher, T., Nenadić, I., Stein, F., Thomas-Odenthal, F., Teutenberg, L., Usemann, P., Dannlowski, U., Hahn, T., Grotegerd, D., Meinert, S., Lencer, R., Tang, Y., Zhang, T., Li, C., Yue, W., Zhang, Y., Yu, X., Zhou, E., Lin, C.-P., Tsai, S.-J., Rodrigue, A. L., Glahn, D., Pearlson, G., Blangero, J., Karuk, A., Pomarol-Clotet, E., Salvador, R., Fuentes-Claramonte, P., Garcia-León, M. Á., Spalletta, G., Piras, F., Vecchio, D., Banaj, N., Cheng, J., Liu, Z., Yang, J., Gonul, A. S., Uslu, O., Burhanoglu, B. B., Uyar Demir, A., Rootes-Murdy, K., Calhoun, V. D., Sim, K., Green, M., Quidé, Y., Chung, Y. C., Kim, W.-S., Sponheim, S. R., Demro, C., Ramsay, I. S., Iasevoli, F., DE Bartolomeis, A., Barone, A., Ciccarelli, M., Brunetti, A., Cocozza, S., Pontillo, G., Tranfa, M., Park, M. T. M., Kirschner, M., Georgiadis, F., Kaiser, S., van Rheenen, T. E., Rossell, S. L., Hughes, M., Woods, W., Carruthers, S. P., Sumner, P., Ringin, E., Spaniel, F., Skoch, A., Tomecek, D., Homan, P., Homan, S., Omlor, W., Cecere, G., Nguyen, D. D., Preda, A., Thomopoulos, S. I., Jahanshad, N., Cui, L.-B., Yao, D., et al. 2024. Neurostructural subgroup in 4291 individuals with schizophrenia identified using the subtype and stage inference algorithm. Nature Communications, 15, 5996.

Joyce, E. M. & Roiser, J. P. 2007. Cognitive heterogeneity in schizophrenia. Current Opinion in Psychiatry, 20.

Kanchanatawan, B., Sriswasdi, S., Thika, S., Sirivichayakul, S., Carvalho, A. F., Geffard, M., Kubera, M. & Maes, M. 2018a. Deficit schizophrenia is a discrete diagnostic category defined by neuro-immune and neurocognitive features: results of supervised machine learning. Metabolic Brain Disease, 33, 1053–1067.

Kanchanatawan, B., Sriswasdi, S., Thika, S., Stoyanov, D., Sirivichayakul, S., Carvalho, A. F., Geffard, M. & Maes, M. 2018b. Towards a new classification of stable phase schizophrenia into major and simple neuro-cognitive psychosis: Results of unsupervised machine learning analysis. Journal of Evaluation in Clinical Practice, 24, 879–891.

Kanchanatawan, B., Thika, S., Sirivichayakul, S., Carvalho, A. F., Geffard, M. & Maes, M. 2018c. In schizophrenia, depression, anxiety, and physiosomatic symptoms are strongly related to psychotic symptoms and excitation, impairments in episodic memory, and increased production of neurotoxic tryptophan catabolites: a multivariate and machine learning study. Neurotoxicity research,33, 641–655.

Kay, S. R., Fiszbein, A. & Opler, L. A. 1987. The positive and negative syndrome scale (PANSS) for schizophrenia. Schizophrenia bulletin,13, 261–276.

Kegel, M. E., Bhat, M., Skogh, E., Samuelsson, M., Lundberg, K., Dahl, M. L., Sellgren, C., Schwieler, L., Engberg, G., Schuppe-Koistinen, I. & Erhardt, S. 2014. Imbalanced kynurenine pathway in schizophrenia. Int J Tryptophan Res,7, 15–22.

Khansari, N., Shakiba, Y. & Mahmoudi, M. 2009. Chronic inflammation and oxidative stress as a major cause of age-related diseases and cancer. Recent patents on inflammation & allergy drug discovery, 3, 73–80.

Kindler, J., Lim, C. K., Weickert, C. S., Boerrigter, D., Galletly, C., Liu, D., Jacobs, K. R., Balzan, R., Bruggemann, J. & O’donnell, M. 2020. Dysregulation of kynurenine metabolism is related to proinflammatory cytokines, attention, and prefrontal cortex volume in schizophrenia. Molecular psychiatry,25, 2860–2872.

Kremen, W. S., Seidman, L. J., Faraone, S. V., Toomey, R. & Tsuang, M. T. 2004. Heterogeneity of schizophrenia: a study of individual neuropsychological profiles. Schizophrenia Research,71, 307–321.

Liu, Y., Tang, Y., Li, C., Tao, H., Yang, X., Zhang, X. & Wang, X. 2020. Altered Expression of Glucocorticoid Receptor and Neuron-Specific Enolase mRNA in Peripheral Blood in First-Episode Schizophrenia and Chronic Schizophrenia. Frontiers in Psychiatry, 11.

Lohr, J. B. 1991. Oxygen radicals and neuropsychiatric illness: some speculations. Archives of general psychiatry, 48, 1097–1106.

Maes, M. 2023a. Major neurocognitive psychosis: a novel schizophrenia endophenotype class that is based on machine learning and resembles Kraepelin’s and Bleuler’s conceptions. Acta Neuro psychiatr,35, 123–137.

Maes, M. 2023b. Major neurocognitive psychosis: a novel schizophrenia endophenotype class that is based on machine learning and resembles Kraepelin’s and Bleuler’s conceptions. Acta Neuropsychiatrica,35, 123–137.

Maes, M. & Kanchanatawan, B. 2022a. In (deficit) schizophrenia, a general cognitive decline partly mediates the effects of neuro-immune and neuro-oxidative toxicity on the symptomatome and quality of life. CNS spectrums, 27, 506–515.

Maes, M. & Kanchanatawan, B. J. C. S. 2022b. In (deficit) schizophrenia, a general cognitive decline partly mediates the effects of neuro-immune and neuro-oxidative toxicity on the symptomatome and quality of life. 27, 506–515.

Maes, M., Moraes, J. B., Congio, A., Bonifacio, K. L., Barbosa, D. S., Vargas, H. O., Michelin, A. P., Carvalho, A. F. & Nunes, S. O. V. 2019a. Development of a novel staging model for affective disorders using partial least squares bootstrapping: effects of lipid-associated antioxidant defenses and neuro-oxidative stress. Molecular Neurobiology, 56, 6626–6644.

Maes, M., Sirivichayakul, S., Kanchanatawan, B. & Carvalho, A. F. 2020a. In schizophrenia, psychomotor retardation is associated with executive and memory impairments, negative and psychotic symptoms, neurotoxic immune products and lower natural IgM to malondialdehyde. The World Journal of Biological Psychiatry, 21, 383–401.

Maes, M., Sirivichayakul, S., Kanchanatawan, B. & Vodjani, A. 2019b. Breakdown of the paracellular tight and adherens junctions in the gut and blood brain barrier and damage to the vascular barrier in patients with deficit schizophrenia. Neurotoxicity research, 36, 306–322.

Maes, M., Sirivichayakul, S., Matsumoto, A. K., Michelin, A. P., De Oliveira Semeão, L., de Lima Pedrão, J. V., Moreira, E. G., Barbosa, D. S., Carvalho, A. F. & Solmi, M. 2020b. Lowered antioxidant defenses and increased oxidative toxicity are hallmarks of deficit schizophrenia: a nomothetic network psychiatry approach. Molecular Neurobiology, 57, 4578–4597.

Maes, M., Sirivichayakul, S., Matsumoto, A. K., Michelin, A. P., de Oliveira Semeão, L., de Lima Pedrão, J. V., Moreira, E. G., Barbosa, D. S., Carvalho, A. F., Solmi, M. & Kanchanatawan, B. 2020c. Lowered Antioxidant Defenses and Increased Oxidative Toxicity Are Hallmarks of Deficit Schizophrenia: a Nomothetic Network Psychiatry Approach. Molecular Neurobiology,57, 4578–4597.

Maes M, Vojdani A, Sirivichayakul S, Barbosa Ds, Kanchanatawan B. Inflammatory and Oxidative Pathways Are New Drug Targets in Multiple Episode Schizophrenia and Leaky Gut, Klebsiella pneumoniae, and C1q Immune Complexes Are Additional Drug Targets in First Episode Schizophrenia. Mol Neurobiol. 2021 Jul;58(7):3319-3334. doi: 10.1007/s12035-021-02343-8. Epub 2021 Mar 6. PMID: 33675500.

Manev, H., Favaron, M., Guidotti, A. & Costa, E. 1989. Delayed increase of Ca2+ influx elicited by glutamate: role in neuronal death. Molecular pharmacology, 36, 106–112.

Masmoudi-Kouki, O., Douiri, S., Hamdi, Y., Kaddour, H., Bahdoudi, S., Vaudry, D., Basille, M., Leprince, J., Fournier, A. & Vaudry, H. 2011. Pituitary adenylate cyclase-activating polypeptide protects astroglial cells against oxidative stress-induced apoptosis. Journal of neuro chemistry, 117, 403–411.

Medina-Hernández, V., Ramos-Loyo, J., Luquin, S., Sánchez, L. F., García-Estrada, J. & Navarro-Ruiz, A. 2007. Increased lipid peroxidation and neuron specific enolase in treatment refractory schizophrenics. J Psychiatr Res, 41, 652–8.

Mondelli, V. & Howes, O. J. P. 2014. Inflammation: its role in schizophrenia and the potential anti-inflammatory effects of antipsychotics. Springer.

Monji, A., Kato, T. & Kanba, S. 2009. Cytokines and schizophrenia: Microglia hypothesis of schizophrenia. Psychiatry and clinical neurosciences, 63, 257–265.

Muller, N. & J Schwarz, M. 2010. The role of immune system in schizophrenia. Current immunology reviews, 6, 213–220.

Neves, D., Salazar, I. L., Almeida, R. D. & Silva, R. M. 2023. Molecular mechanisms of ischemia and glutamate excitotoxicity. Life Sciences, 121814.

Overall, J. E. & Gorham, D. R. 1962. The brief psychiatric rating scale. Psychological Reports, 10, 799–812.

Palmqvist, S., Janelidze, S., Quiroz, Y. T., Zetterberg, H., Lopera, F., Stomrud, E., Su, Y., Chen, Y., Serrano, G. E. & Leuzy, A. 2020. Discriminative accuracy of plasma phospho-tau217 for Alzheimer disease vs other neurodegenerative disorders. Jama,324, 772–781.

Palumbo, B., Sabalich, I., Tranfaglia, C. & Lucilla Parnetti, M. J. F. N. 2008. Cerebrospinal fluid neuron-specific enolase: a further marker of Alzheimer’s disease? 23, 93.

Pavan, T., Aleman-Gomez, Y., Jenni, R., Steullet, P., Schilliger, Z., Dwir, D., Cleusix, M., Alameda, L., Do, K. Q. & Conus, P. 2024. White Matter Microstructure Alterations and Their Link to Symptomatology in Early Psychosis and Schizophrenia. medRxiv, 2024-02.

Pedraz-Petrozzi, B., Elyamany, O., Rummel, C. & Mulert, C. 2020. Effects of inflammation on the kynurenine pathway in schizophrenia - a systematic review. J Neuroinflammation, 17, 56.

Popov, P., Chen, C., Al-Hakeim, H. K., Al-Musawi, A. F., Al-Dujaili, A. H., Stoyanov, D. & Maes, M. 2024. The novel schizophrenia subgroup “major neurocognitive psychosis” is validated as a distinct class through the analysis of immune-linked neurotoxicity biomarkers and neurocognitive deficits. Brain Behav Immun Health, 40, 100842.

Roomruangwong, C., Noto, C., Kanchanatawan, B., Anderson, G., Kubera, M., Carvalho, A. F. & Maes, M. 2020. The Role of Aberrations in the Immune-Inflammatory Response System (IRS) and the Compensatory Immune-Regulatory Reflex System (CIRS) in Different Phenotypes of Schizophrenia: the Irs-CIRS Theory of Schizophrenia. Mol Neurobiol, 57, 778–797.

Rothermundt, M., Ahn, J. N. & Jorgens, S. 2009. S100B in schizophrenia: an update. Gen Physiol Biophys, 28, F76–81.

Sas, K., Szabó, E. & Vécsei, L. 2018. Mitochondria, Oxidative Stress and the Kynurenine System, with a Focus on Ageing and Neuroprotection. Molecules, 23.

Schönknecht, P., Hempel, A., Hunt, A., Seidl, U., Volkmann, M., Pantel, J. & Schröder, J. 2003. Cerebrospinal fluid tau protein levels in schizophrenia. European archives of psychiatry and clinical neuroscience, 253, 100–102.

Schwarcz, R., Bruno, J. P., Muchowski, P. J. & Wu, H. Q. 2012. Kynurenines in the mammalian brain: when physiology meets pathology. Nat Rev Neurosci, 13, 465–77.

Silvestro, S., Raffaele, I., Quartarone, A. & Mazzon, E. 2024. Innovative Insights into Traumatic Brain Injuries: Biomarkers and New Pharmacological Targets. International Journal of Molecular Sciences, 25, 2372.

Sirivichayakul, S., Kanchanatawan, B., Thika, S., Carvalho, A. F. & Maes, M. 2019a. Eotaxin, an endogenous cognitive deteriorating chemokine (ECDC), is a major contributor to cognitive decline in normal people and to executive, memory, and sustained attention deficits, formal thought disorders, and psychopathology in schizophrenia patients. Neurotoxicity research, 35, 122–138.

Sirivichayakul, S., Kanchanatawan, B., Thika, S., Carvalho, A. F. & Maes, M. 2019b. A new schizophrenia model: immune activation is associated with the induction of different neurotoxic products which together determine memory impairments and schizophrenia symptom dimensions. CNS Neurological Disorders-Drug Targets,18, 124–140.

Skorobogatov, K., Autier, V., Foiselle, M., Richard, J. R., Boukouaci, W., Wu, C. L., Raynal, S., Carbonne, C., Laukens, K., Meysman, P., Coppens, V., Le Corvoisier, P., Barau, C., De Picker, L., Morrens, M., Tamouza, R. & Leboyer, M. 2023. Kynurenine pathway abnormalities are state-specific but not diagnosis-specific in schizophrenia and bipolar disorder. Brain Behav Immun Health, 27, 100584.

Steiner, J., Bielau, H., Bernstein, H. G., Bogerts, B. & Wunderlich, M. T. 2006. Increased cerebrospinal fluid and serum levels of S100B in first-onset schizophrenia are not related to a degenerative release of glial fibrillar acidic protein, myelin basic protein and neurone-specific enolase from glia or neurones. J Neurol Neurosurg Psychiatry,77, 1284–7.

Steiner, J., Walter, M., Wunderlich, M. T., Bernstein, H.-G., Panteli, B., Brauner, M., Jacobs, R., Gos, T., Rothermundt, M. & Bogerts, B. 2009. A new pathophysiological aspect of S100B in schizophrenia: potential regulation of S100B by its scavenger soluble RAGE. Biological psychiatry, 65, 1107–1110.

Wang, K. K., Zhang, Z. & Moghieb, A. 2015. Biomarkers for CNS injury and regeneration. Neural Regeneration. Elsevier.

Yang, Q., Zhang, Y., Yang, K., Niu, Y., Fan, F., Chen, S., Luo, X., Tan, S., Wang, Z., Tong, J., Yang, F., Li, C. R. & Tan, Y. 2022. Associations of the serum kynurenine pathway metabolites with P50 auditory gating in non-smoking patients with first-episode schizophrenia. Front Psychiatry, 13, 1036421.

Yu, C.-P., Pan, Z.-Z. & Luo, D.-Y. 2016. TDO as a therapeutic target in brain diseases. Metabolic brain disease, 31, 737–747.

Zhang, Z., Zhang, S., Fu, P., Zhang, Z., Lin, K., Ko, J. K.-S. & Yung, K. K.-L. 2019. Roles of glutamate receptors in Parkinson’s disease. International journal of molecular sciences, 20, 4391.

