## Supplementary material for "Brain injury biomarkers in major and simple neurocognitive psychosis: association with tryptophan catabolites": ESF, Table 1. ESF, Table 2.

**Electronic supplementary file (ESF)**

**ESF, Table 1.** Assessment of the brain injury biomarkers in schizophrenia patients and healthy controls.

| Parameter | Healthy Control | Schizophrenia | F | df | p |
| --- | --- | --- | --- | --- | --- |
| S100B (pg/ml) | 14.95 (2.65) | 20.09(1.87) | 0.18 | 1/174 | 0.675 |
| pTau217 (pg/ml) | 3.10 (0.20) | 3.81 (0.14) | 7.41 | 1/174 | 0.007 |
| Nestin (pg/ml) | 71.93 (5.29) | 70.05 (3.73) | 0.08 | 1/174 | 0.772 |
| NSE (pg/ml) | 0.48 (0.04) | 0.82 (0.03) | 64.34 | 1/174 | <0.001 |
| GFAP (pg/ml) | 1.32 (0.51) | 4.15 (0.36) | 86.40 | 1/174 | <0.001 |
| BII (z score) | -1.754 (0.303) | 0.877 (0.213) | 50.039 | 1/174 | <0.001 |

All data are shown as estimated marginal mean (SE). All results of GLM analysis (adjusted for sex, age, smoking and BMI)

GFAP: Glial fibrillary acidic protein, S100B: S100 calcium-binding protein B, NSE: Neuron-Specific Enolase, pTau217: **phosphorylated tau protein at amino acid 217. BII: brain injury index**

**ESF, Table 2.** Intercorrelations among brain injury biomarkers and tryptophan and tryptophan catabolites in patients with schizophrenia.

| Parameters | pTau217 | NSE | GFAP | BII |
| --- | --- | --- | --- | --- |
| Tryptophan | 0.092 (0.319) | -0.162 (0.077) | -0.151 (0.100) | -0.125 (0.175) |
| Kynurenine | -0.106 (0.25) | **-0.323 (<0.001)** | -0.189 (0.038) | -0.169 (0.064) |
| 3-hydroxy-kynurenine | -0.134 (0.145) | -0.137 (0.135) | 0.001 (0.992) | -0.111 (0.229) |
| Kynurenine acid (KA) | -0.174 (0.057) | **-0.338 (<0.001)** | **-0.194 (0.034)** | **-0.290 (0.001)** |
| Anthranilic acid | 0.013 (0.886) | 0.124 (0.179) | 0.039 (0.675) | 0.108 (0.242) |
| 3-hydroxy-anthranilic acid | -0.131 (0.154) | -0.104 (0.259) | **-0.216 (0.018)** | **-0.189 (0.039)** |
| Quinolinic acid (QA) | **0.187 (0.041)** | **0.239 (0.009)** | **0.202 (0.027)** | **0.212 (0.02)** |
| zQA-zKA | **0.296 (0.001)** | **0.401 (<0.001)** | **0.301 (0.001)** | **0.356 (<0.001)** |

GFAP: Glial fibrillary acidic protein, NSE: Neuron-Specific Enolase, pTau217: **phosphorylated tau protein at amino acid 217. BII: brain injury index;** zQA-zKA: index for the QA / KA ratio.
